# Federated Prediction Models and External Validation for Radiotherapy Outcomes in Oropharyngeal Cancer using F.A.I.R. Clinical and Radiomics Data

**DOI:** 10.1101/2025.01.29.25321214

**Authors:** Varsha Gouthamchand, Benedetta Gottardelli, Gauri Kulkarni, Umesh B Sherkhane, Joshi Hogenboom, Rajamenakshi Subramanian, Ashish Kumar Jha, Sneha Mithun, Nilendu C Purandare, Jai Prakash Agarwal, Krithikaa Sekar, G. Lohith, Sarbani G Laskar, Shwetabh Sinha, Frank JP Hoebers, Venkatesh Rangarajan, Gaur Sunder, Andre Dekker, Johan van Soest, Leonard Wee

## Abstract

Development and validation of outcome prediction models in multi-centric cancer datasets are essential to ensure their applicability and accuracy across diverse populations. This study addresses the challenges of model generalizability in Head and Neck cancer research by utilizing combined clinical and radiomics data from centers in India and the Netherlands, following Findable-Accessible-Interoperable-Reusable (F.A.I.R.) data principles. We use Vantage6, a federated learning software infrastructure that implements the Personal Health Train (PHT) paradigm to ensure data privacy and security during collaborative research. Correlation-based Feature Selection (CFS) and LASSO regularized Cox regression were used to identify key features in training Cox proportional hazards models to predict Overall Survival (OS), Distant Metastasis (DM), and Locoregional Recurrence (LRR) in six datasets totaling 1131 oropharyngeal cancer patients. Our results highlight the efficacy of federated learning in providing a secure environment for multi-centric cancer research, enabling the development and validation of predictive models while upholding patient data confidentiality.

## Introduction

Head and Neck cancers (H&N) in India account for a significant fraction of the total cancer burden, including more than 30% of all cancer cases globally, whereas in the Netherlands, the percentage is just above 2%. In the Netherlands, the 5-year survival rate is 57%, while in India it is approximately 32% [1, 2]. Differences in mortality outcome may be attributed to differences in genetic features, Human Papilloma Virus (HPV) infection, nicotine usage, alcohol consumption, stage at presentation, distribution over H&N anatomical sites, and dietary patterns, among other risk factors [3]. The high incidence rates in India, coupled with delayed reporting, suggest challenges in early detection and access to medical care.

Relatively new and advanced treatment options, such as proton beam therapy and magnetic resonance imaging-guided adaptive radiation therapy, complicate medical decision-making. Consequently, there is a growing need for personalized clinical decision support systems that can enhance treatment, whilst considering individual preferences. The **TRAIN (personal health Train for RAdiation oncology in India and the Netherlands) project** [4], an Indo-Dutch collaborative initiative, aims to develop Decision Support Systems (DSS) using multi-centric prediction models that integrate data from India and the Netherlands. This collaboration seeks to enhance treatment outcomes for HNSCC patients through improved clinical decision-making.

When developing multi-centric outcome prediction models in cancer research, many external validations are needed before clinical implementation, but this is often overlooked [5, 6]. Validating a model’s utility, accuracy, and discriminatory power is necessary to prove it is safe to be used in a specific scenario [7]. Especially when relying on limited or single-center datasets, models may produce biased results that do not generalize well with unseen data [8].

Harnessing real-world clinical data at scale poses several challenges, including syntactic interoperability issues and patient privacy concerns. Federated Learning approaches, manifested by the Personal Health Train (PHT), have emerged as a viable solution to address privacy related challenges [9, 10]. This has the potential to empower data owners with complete control over their data, a concept gaining momentum over the recent years [11–15]. Implementing such approaches hinges on adhering to F.A.I.R. principles (Findable, Accessible, Interoperable, and Reusable) [16, 17] data principles. Linked Data, with its focus on interconnecting data through machine-readable URIs [18], provide a framework aligning with the F.A.I.R. principles.

Building upon our previous work on F.A.I.R.-ification of structured clinical data [19], this work extends the approach by integrating clinical data, radiomics features, and Digital Imaging and Communications in Medicine (DICOM) headers - as Linked Data. The integrated data from multiple centers are presented through an interactive visualization dashboard for initial federated data exploration, thereby laying a foundation for survival analysis aimed at personalized healthcare delivery.

Radiomics, standardized by the Image Biomarker Standardization Initiative (IBSI) [20], are quantitative biomarkers extracted from medical images [21]. It holds tremendous promise for improving outcome predictions, especially by addressing the intricate intra-tumoral heterogeneity and anatomical complexities prevalent in H&N cancers. Consequently, numerous studies have emerged to explore radiomics’ applications in H&N cancer diagnosis [22–29]. However, the high dimensionality of radiomics data, with hundreds to thousands of variables extracted, poses a significant challenge. Effective feature selection is essential to manage this dimensionality, as demonstrated in our previous collaborative research on multi-centered distributed radiomics for non-small cell lung cancer [30].

The primary objective of this paper is to apply the previously developed federated feature selection pipeline to Linked Data from the TRAIN consortium centers, following the initial data exploration. The identified subsets were employed in federated multivariate Cox Proportional Hazards (CoxPH) models to predict Overall Survival (OS), Distant Metastasis (DM), and Loco-Regional Recurrence (LRR). This study also aims to external validate these models to demonstrate the viability and robustness of federated multi-centric analysis.

## Methods

### Consortium agreement, Data Collection, and Approval for Data Protection

This is a retrospective multicenter federated data study conducted under the consortium “TRAIN” (ClinicalTrials.gov Identifier NCT04655469) and established through a signed Consortium Research Agreement between MAASTRO Clinic, Healthcare Global (HCG), and Tata Memorial Hospital (TMH). Approval for re-use of standard-of-care (non-investigational) observational patient data in each partner institution was granted by their respective ethics review boards - waiving patient informed consent for the study.

All the methods and findings presented in this paper adhere to all relevant ethical guidelines and regulations governing the treatment of human subjects, ensuring compliance with ethical norms throughout the research and manuscript preparation.

### Data preparation

The data preparation process included integrating clinical, radiomics, and DICOM header information at each participating centers. Each center installed the Flyover tool (introduced in our previous work [19]) for data conversion. For this study, an upgraded version of the tool was employed, featuring new components for handling radiomics and DICOM headers, in addition to converting structured data (**Figure 1**). The tool comprises two Docker packages functioning independently: one for data processing and conversion, and the other package for RDF-store. After data conversion, the first package can be deactivated while the RDF-store continues to operate, optimizing computational memory. More details on the tool’s usage can be found in the public GitHub repository (https://github.com/MaastrichtU-CDS/Flyover).

**Figure 1.**
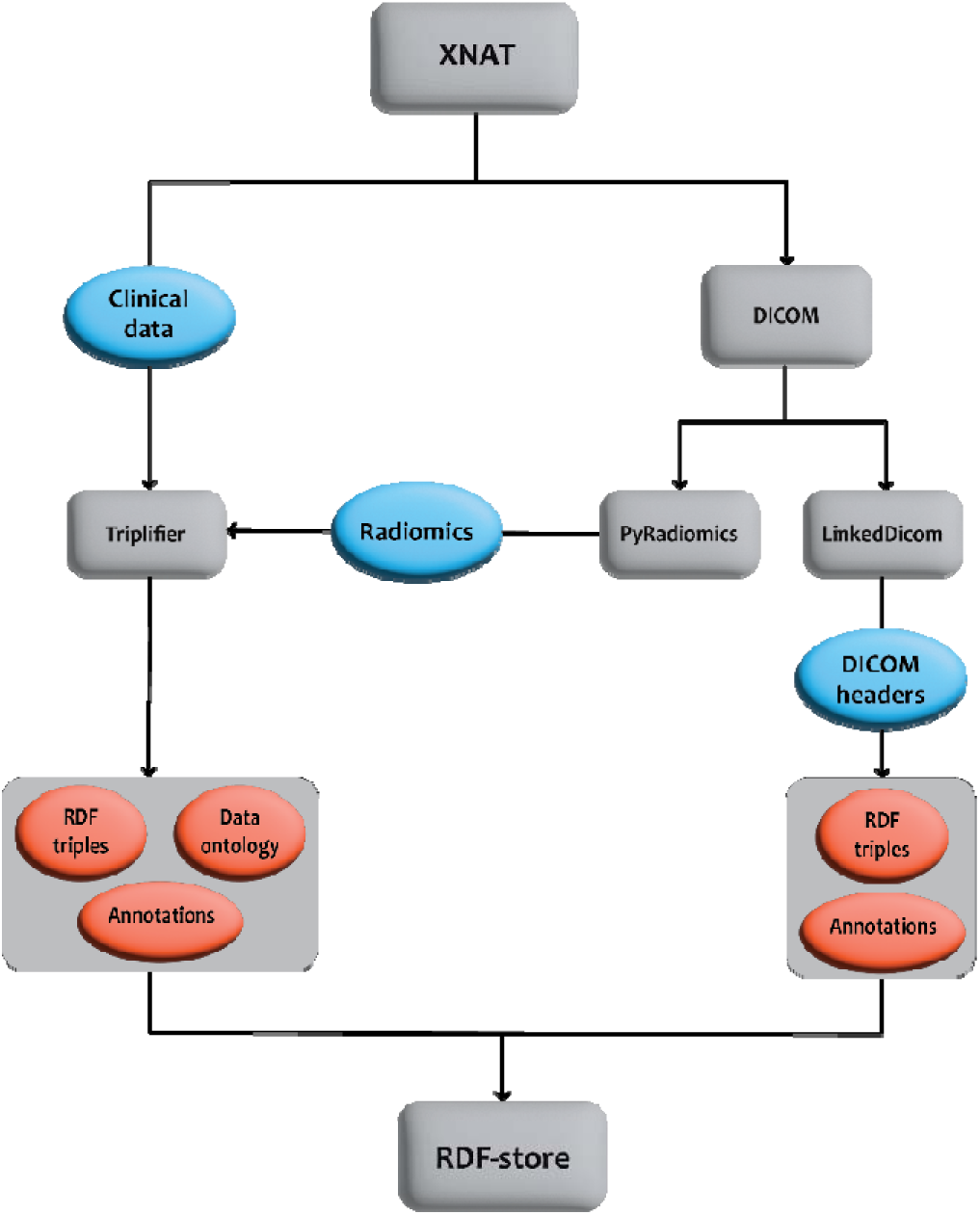
Illustration of the F.A.I.R. conversion workflow. Blue circles represent the data undergoing conversion, while red circles represent the final RDF-triples utilized in the analysis.

### Infrastructure topology

For our federated-learning infrastructure, we rely on Vantage6 (v3.7.3) [31], an open-source framework adhering to the PHT approach. Detailed insights can be found in the associated paper. The three public datasets - **HN1** [32], **HEAD-NECK** [33], **OPC-RADIOMICS** [34] were hosted on cloud-based virtual machines in Europe (Ubuntu OS) maintained and controlled by the authors, while three proprietary datasets were securely stored within their respective institutions: two in India – **HCG and TMH** (Windows OS) and one in the Netherlands – **HN3** (CentOS). These distributed hosts were connected to a centralized Vantage6 trusted server, hosted by Centre for Development of Advanced Computing (CDAC) in Pune, India. The architecture is illustrated in **Figure 2**.

**Figure 2.**
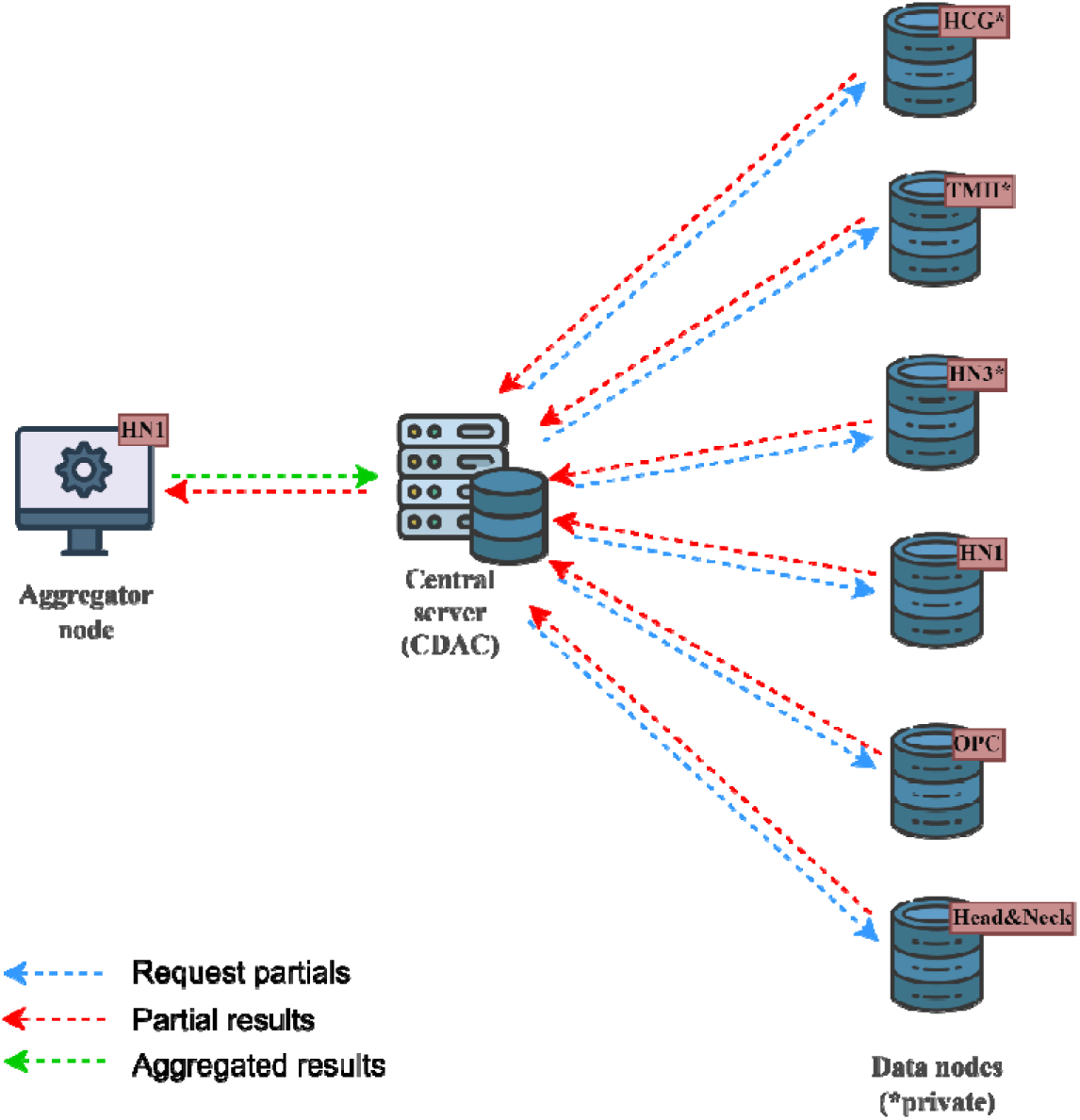
Schematic illustration of the Vantage6 infrastructure used in this work.

### Federated exploration of F.A.I.R. annotated Clinical, DICOM, and Radiomics data

We developed an interactive dashboard for initial data exploration and quality control, similar to our previous work [19]. This involved distributing federated SPARQL queries and cross-referencing the Linked Data in each of the six data nodes (or data nodes). The HN1 node acted as the aggregator, consolidating partial results from the other nodes, before transmitting the final aggregated results to the central server. The outcomes were visualized for the researcher in the interactive dashboard (**Figure 3**).

**Figure 3.**
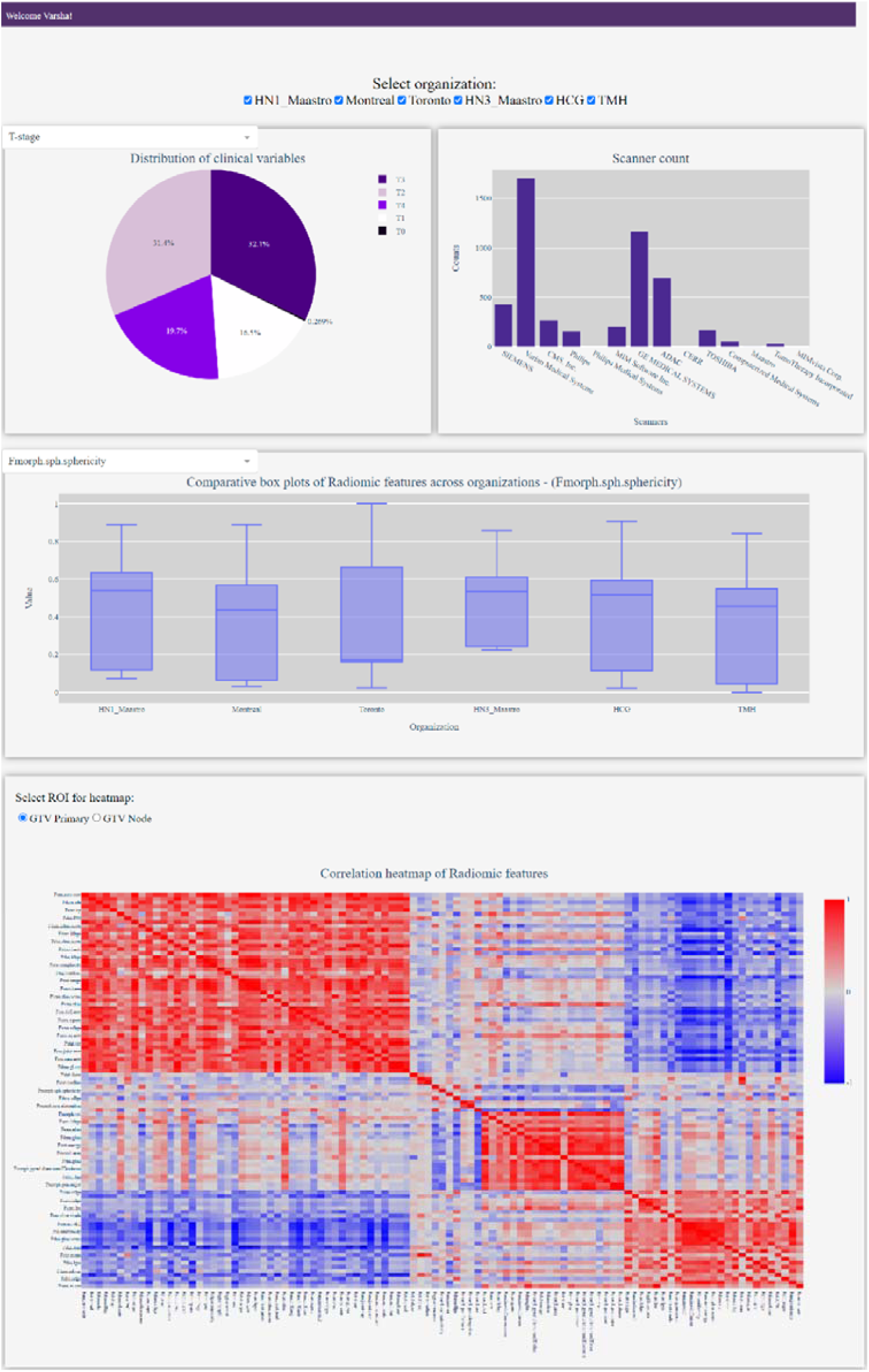
Screen capture of the prototype dashboard.

### Federated Feature Selection

For feature selection, we utilized strategies introduced in the previous study [30]: Correlation-based Feature Selection (CFS) and the LASSO regularized Cox regression model, both implemented within the Vantage6 framework. Clinical variables were handpicked, including T-staging, N-staging, HPV status and Chemotherapy. T-staging was split into two categorical predictors: T3-or-lower and T3-or-higher. Similarly for N-staging as N1-or-lower and N1-or-higher. Chemotherapy was treated as a categorical variable indicating whether it was administered or not. Due to a significant amount of missing data for HPV status, an additional predictor, HPV-unknown, was included alongside the HPV-positive and HPV-negative categories.

The clinical variables were processed with LASSO to determine the feature rankings for subsequent Cox regression analysis. To manage the high dimensionality of radiomics features, CFS was used to reduce the feature pool, followed by an additional CFS iteration to separate highly correlated features in the first iteration (correlation coefficient > 0.70) into batches. These refined subsets were then processed with LASSO. This segregation aimed to mitigate the computational burden associated with LASSO when handling highly correlated variables.

The optimal feature set was determined using Leave-One-Out Cross-Validation (LOOCV) - training CoxPH model on two data nodes and validating on the third across the training cohorts. The feature subset with the highest Harrell’s concordance index (C-index) during LOOCV was selected for the final model training.

### Federated model training and external validation

A series of CoxPH models were trained using selected optimal feature sets to predict OS, DM and LRR outcomes for oropharyngeal cancer patients. The models integrated clinical and radiomics data into four model frameworks: a clinical-only model (C-Model), a radiomics model using primary Gross Tumor Volume (GTV) features (RP-Model), a radiomics model using nodal GTV features (RN-Model), and an integrated model using the linear predictors from the three aforementioned models (CR-Model). Feature selection (for C-Model, RP-Model and RN-Model) and model training for each framework were executed on a combination of designated training cohorts within each loop (**Figure 4**) for each outcome, followed by individual validation on the validation cohorts. The resulting C-indices were averaged to achieve a global C-index. This comprehensive approach enabled broad analysis and insights from the patient groups. The related model code is available in the public GitHub repository (https://github.com/MaastrichtU-CDS/TRAIN-DCR-tools).

**Figure 4.**
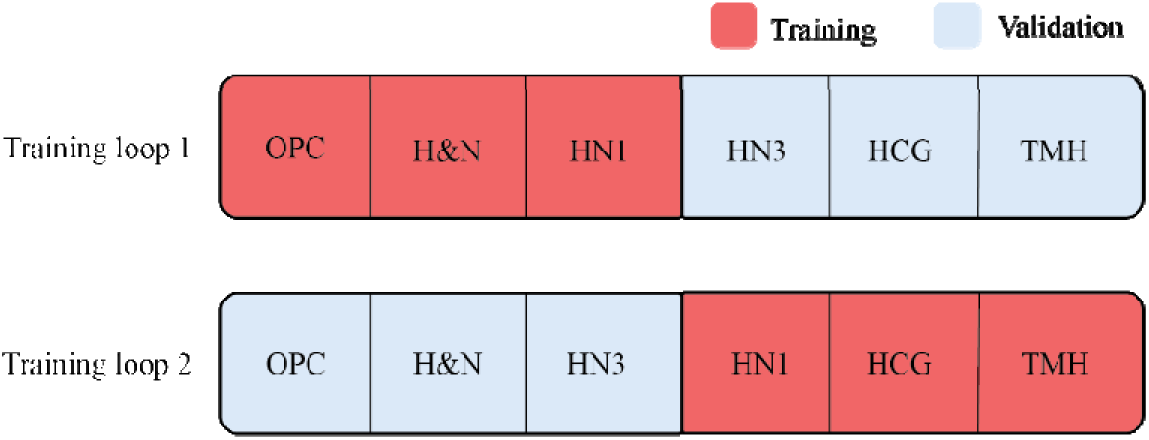
Training and validation loops.

## Results

### Data description

The six datasets comprised of 1814 patients, with a mean age of 60.8 years. For this study, we used a subset of 1131 oropharyngeal cancer patients, the general characteristics of which are summarized in **Supplementary Table S1**. In addition to clinical parameters, which included long-term follow-up information, we also obtained radiotherapy planning Computed Tomography (CT) images (in DICOM format). The primary and nodal gross tumor volume (GTV) were manually delineated by expert radiation oncologists (as RTSTRUCT) for each dataset. The CT images and clinical data for the three public datasets can be openly accessed in The Cancer Imaging Archive (TCIA) [35]. Further description of the datasets can be found in the Data Availability section.

### Annotated Linked Data

The Resource Description Framework storage (RDF-store) instances running on each node in the federated infrastructure serve as a comprehensive repository, housing triples from three categories - clinical, radiomics, and DICOM headers - and their integration is executed manually through SPARQL insert queries. This process entails matching patient IDs across records within the clinical and radiomics graphs and aligning patient IDs and SOP (Service Object Pair) instance UIDs across objects in the radiomics and DICOM graphs. As a result of this, these graphs are interlinked within a coherent semantic data model, thus forming the Linked Data (**Figure 5**). This enriched model, augmented with contextual knowledge, not only enhances machine readability but also positions itself as an ideal candidate for deployment in federated learning scenarios.

**Figure 5.**
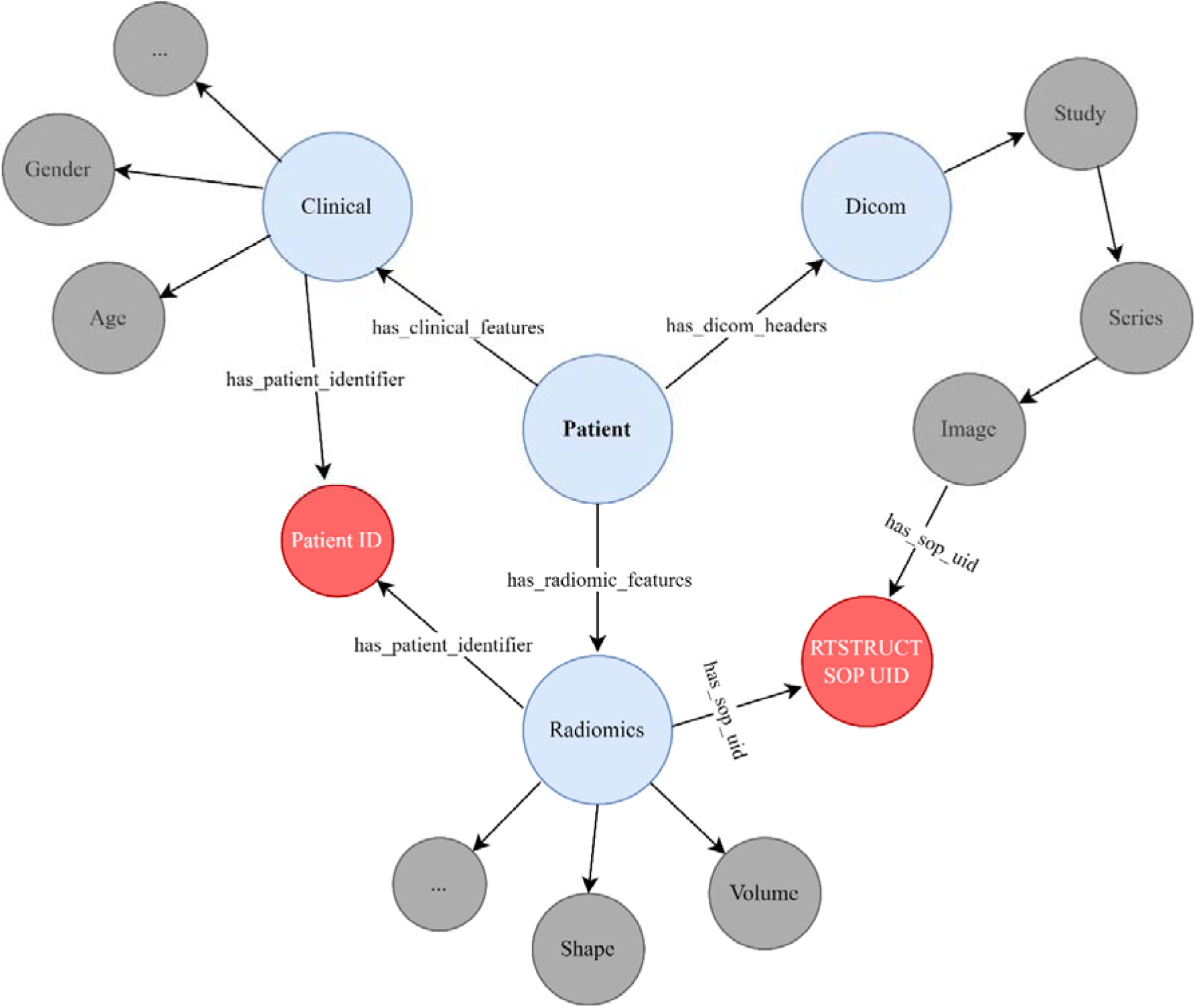
Clinical, Radiomics and DICOM headers interconnected as Linked Data

### Data exploration dashboard

**Figure 3** provides a snapshot of the distributed interactive dashboard, offering researchers a comprehensive overview of the data available across the geographically dispersed F.A.I.R. data nodes. This use-case dashboard extends beyond our prior work, which displayed only clinical parameters. In this new version, users can also explore the distribution of each radiomics feature across the nodes and view correlations among radiomics via a distributed correlation heat map. Additionally, the dashboard provides the option to view scanner information from various institutions, extracted directly from the DICOM headers.

### Feature selection, multivariate Cox models and external validation

The optimal subsets of variables selected for the final CoxPH analysis following the federated feature selection methods, are presented in **Table 1**, **Table 2, and Table 3** for the C-Model, RP-Model and RN-Model respectively. The CR-Model uses the linear predictors derived from these three models as its variables.

**Table 1.** Optimal feature subsets selected for C-Model.

| C-Model | Loop 1 | Loop 2 |
| --- | --- | --- |
| **OS** | T3-or-higher<br>HPV negative<br>HPV positive<br>N1-or-lower<br>Chemotherapy | HPV negative<br>HPV positive<br>N1 or lower<br>Chemotherapy |
| **DM** | T3-or-higher<br>HPV negative<br>N1-or-lower<br>Chemotherapy | HPV negative<br>HPV positive<br>N1 or lower<br>Chemotherapy |
| **LRR** | T3-or-higher<br>HPV negative<br>HPV positive<br>Chemotherapy | HPV negative<br>HPV positive<br>N1 or lower<br>Chemotherapy |

**Table 2.** Optimal feature subsets selected for RP-Model.

| RP-Model | Loop 1 | Loop 2 |
| --- | --- | --- |
| OS | Fszm sze<br>Fmorph sph sphericity<br>Fmorph av | Fmorph pca elongation<br>Fcm inv var |
| DM | Fszm zlnu norm<br>Fmorph sph sphericity<br>Fmorph av | Fcm sum energy<br>Fmorph sph sphericity |
| LRR | Fszm zlnu norm<br>Fcm inv var<br>Fmorph sph sphericity<br>Fmorph av<br>Fdzm sdlge | Fcm corr<br>Frlm lrlge |

**Table 3.** Optimal feature subsets selected for RN-Model.

| RN-Model | Loop 1 | Loop 2 |
| --- | --- | --- |
| OS | Fdzm sde<br>Frlm lrlge | Fcm corr<br>Fszm lgze<br>Fdzm lgze<br>Fstat skew |
| DM | Fszm sze<br>Fcm info corr 1 | Fmorph sph sphericity<br>Frlm rlnu norm |
| LRR | Fstat skew<br>Frlm glnu norm<br>Frlm srlge | Fdzm sdlge<br>Fdzm sde<br>Fcm info corr 1 |

The averaged global C-indices of the model outcomes validated across the validation cohorts in both training loops, are depicted in **Figure 6**. Across both loops, the C-Model emerged as the highest performing model for OS, maintaining a consistent global average C-index of 0.62. Similarly for DM, the C-Model exhibited higher performance with average C-indices of 0.64 and 0.63 for Loop 1 and Loop 2, respectively. However, the results for LRR displayed variability. In Loop 1, the RN-Model showcased the highest performance with a C-index of 0.64, while in Loop 2, the CR-Model outperformed others with a C-index of 0.64. For a detailed overview of the results of the trained CoxPH models in each loop, including the hazard ratio (HR) estimates and C-indices of the individual validation cohorts, please refer to the Supplementary materials.

**Figure 6.**
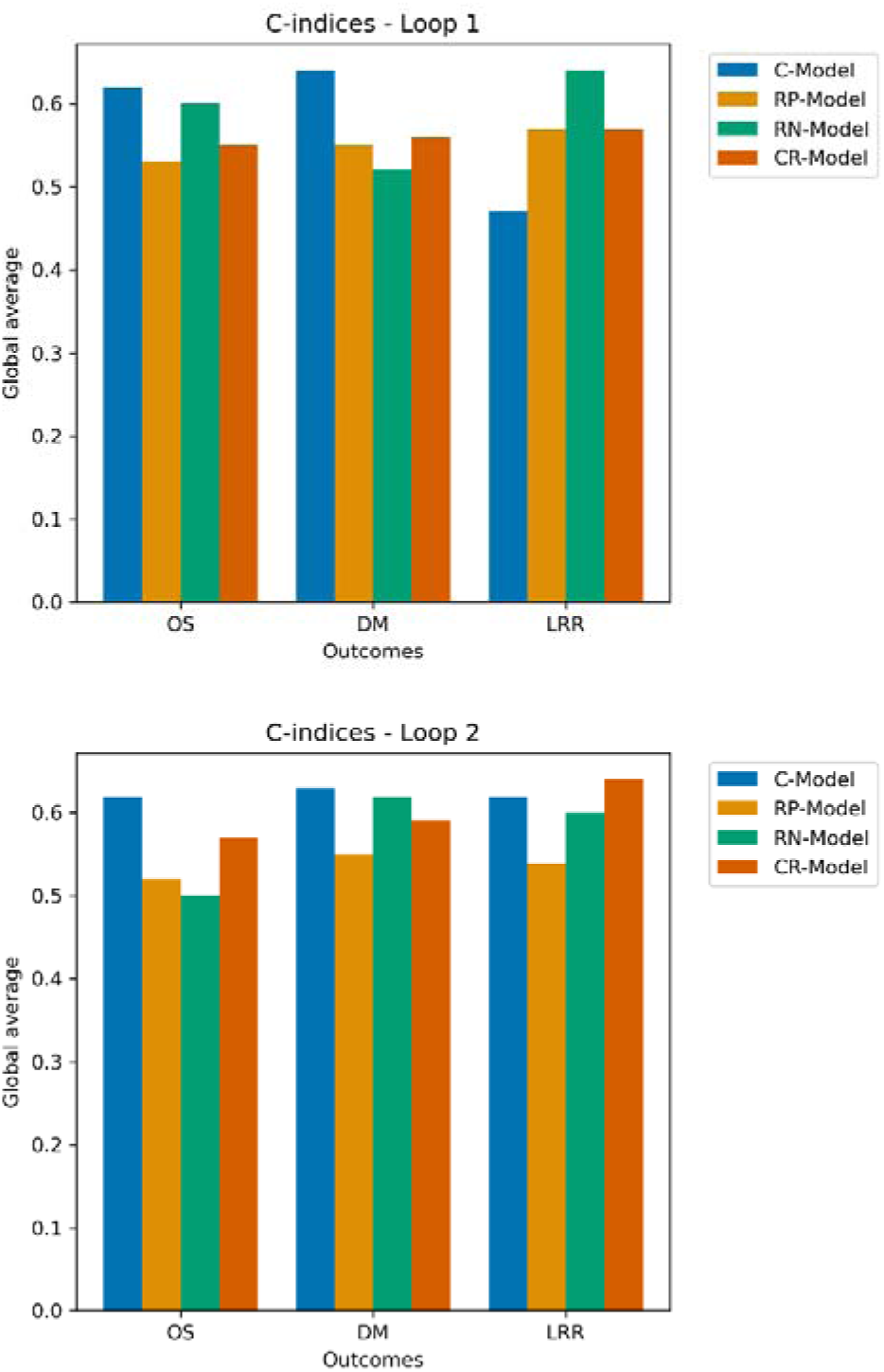
Global averaged C-indices for Loop 1 and Loop 2

## Discussion

In this paper, we extend the methodologies previously applied to the public datasets and the private Maastro HN3 dataset by incorporating private datasets from India. Our objective is to enhance these methods and assess the robustness of the survival models when validated against new, unseen data that feature distinct patient characteristics. We also investigate the integration of diverse information sources by linking three domains of clinical knowledge into a unified data model, leveraging the capabilities of Linked Data. A related study by Kalendralis et al. in 2020 [36] introduced a workflow to transform clinical, radiomics, and DICOM data into RDF format. Our tool aims to enhance data conversion and interlinking methodologies, for a platform-independent integration process. This refinement ensures ease of deployment for non-technical stakeholders while enhancing overall efficiency. Furthermore, by incorporating feature selection and reduction techniques, we aim to demonstrate a comprehensive use-case for applying federated learning tools in a clinical context.

The healthcare landscape has witnessed an exponential surge in the accumulation of electronic patient records within various healthcare institutions. Professionals in the healthcare domain are eager to leverage this wealth of data for research, with the aim of improving personalized patient care. However, it is widely acknowledged that data sourced from a single institution often falls short in adequately addressing complex research questions or in training accurate prediction models. Against this backdrop, federated learning approaches such as the PHT, are gaining significant traction within the healthcare domain. This study not only offers a compelling and clinically relevant use-case of federated learning’s potential in advancing clinical research but also highlights its diverse applications in cancer care.

While other formats like Azure Data Cubes and OMOP-CDM can be used for F.A.I.R. compliance, RDF is particularly advantageous, which is attributed to several compelling reasons. Owing to its graphical nature, RDF significantly streamlines data exploration and integration when compared to alternative data storage formats. Technologies such as RDFS (RDF Schema) and OWL (Web Ontology Language) enhance the semantic richness of data, allowing for more precise data relationships and inferencing capabilities, which are crucial for advanced data analysis, interoperability and reusability. Moreover, RDF’s design inherently supports linking data from disparate sources, making it easier to integrate and query data across multiple systems and domains [37].

Analyzing the performance metrics across the four models provides significant insights. C-Model consistently demonstrated robust predictive power for overall survival (OS), achieving a global average C-index of 0.62. This highlights the continued importance of traditional clinical parameters as reliable indicators of overall prognosis, a fact supported by their widespread use in clinical practice and randomized trials. For distant metastasis (DM), the C-Model also excelled, with average C-indices of 0.64 and 0.63 in Loop 1 and Loop 2, respectively. This underlines the clinical parameters’ predictive strength across various survival endpoints, which is crucial for comprehensive patient management.

However, the results for the locoregional recurrence (LRR) showcased differences in both the loops. In Loop 1 (validated with the Indian and Dutch datasets), RN-Model was the most effective, with a C-index of 0.64, suggesting that radiomics features related to nodal characteristics are potent predictors of LRR. In Loop 2 (validated with the Canadian and Dutch data), the combined CR-Model showed the highest performance, also with a C-index of 0.64. This variation highlights the potential of integrating radiomics with clinical data to enhance predictive accuracy, particularly in complex cases where traditional methods may falter. These findings emphasize the importance of model selection based on the specific survival outcome of interest. They also suggest that while clinical models are generally effective, incorporating radiomics can potentially offer improvements, especially in predicting more localized disease processes such as LRR.

We carried out model training and validation in two separate loops, dividing the datasets to separately analyze public and private datasets within a federated framework. This allowed us to gain insights into the differences between data and modelling results from developed western regions like the Netherlands and that from developing countries, such as India. Additionally, some cases like the categorical predictor “Chemotherapy” in our clinical model consistently resulted in better prognosis. However, as is common in predictive modeling, this may be influenced by potential biases, such as the tendency for patients with favorable prognosis to receive concurrent chemotherapy. At this point, we remain uncertain whether this was indeed a confounding variable in our model’s results.

This study has a few limitations that we must acknowledge. First, we encountered challenges due to the heterogeneous nature of data sourced from six different datasets. The variability across organizations, as highlighted by the box plots in the Supplementary section, reflects the influence of differing imaging protocols and patient demographics. For example, in loop 1, the selected feature *Fmorph.sph.sphericity* for prediction of DM displayed a relatively consistent range across HN1 and HEAD-NECK, whereas OPC exhibited wider variability. The reasons could include potential heterogeneity in tumor shape characteristics, with a trend towards less spherical tumors among patients from OPC or varying tumor contouring guidelines.

Similarly, features like *Fcm.sum.energy* selected in loop 2 for DM showed variations in central tendencies and spread across datasets, indicating differences in texture and intensity captured in the images. These discrepancies could be attributed to the lack of standardized imaging protocols, varying scanners, image acquisition parameters, and reconstruction settings used by different organizations. Such variations impact the robustness and reproducibility of the extracted radiomic features.

Future work could address this issue by extracting imaging parameters, such as slice thickness and Kilovoltage peak (kVp), along with other DICOM headers using an updated version of the LinkedDicom tool. Adding these in a federated dashboard will provide an overview of different imaging parameters, facilitating better handling of multi-center studies. We also propose exploring image-based harmonization techniques, including deep learning approaches like Generative Adversarial Networks (GANs) [38], to translate images across different acquisition settings. Alternatively, feature harmonization methods like ComBat [39–42] can adjust features to account for batch effects and other sources of variability.

Secondly, while the federated Cox regression model does not share sensitive data, it does share a list of unique event times for aggregation from each center. Although not typically considered re-identifiable, there is a risk of deducing event times for the involved patients, particularly with fewer datasets or with the involvement of public datasets. Kairouz et al [43] discusses additional problems and challenges associated with federated learning and proposes solutions, such as enhancing privacy and preventing model inversion using technologies like distributed homomorphic encryption or distributed differential privacy. These solutions are areas we plan to explore in our future work.

Our methods employed the C-index as the performance score metric, but we did not calculate confidence intervals for the C-indices, which could be included in future predictive modeling efforts. Furthermore, including additional accuracy measures such as Brier scores or Root Mean Squared Error (RMSE) could offer further insights into the overall performance of our models.

Finally, calculating the global baseline hazard function to represent the hazard rate for the reference group (when all covariates are at their reference levels) in multi-centric cancer data is essential. To our knowledge, this has not been investigated in a federated context. Establishing a benchmark reference curve for the participating data sources would provide invaluable insights into the impact of various factors on event hazard rates over time. Additionally, validating the assumptions of survival models is a crucial step that we plan to explore in our upcoming work. This can be achieved by plotting Schoenfeld residuals against time and other covariates. Schoenfeld residuals, which are the differences between predicted and observed covariate values for patients at risk, are independent of time [44]. Plotting them against time will enable us to observe and analyze trends and patterns of the included data in a multi-centric collaboration.

## Conclusions

This study effectively utilized federated learning techniques to develop and externally validate predictive models for H&N cancer outcomes using a mix of clinical and radiomics data from multiple centers. By adopting the PHT approach, we ensured that data analysis was secure and privacy conscious. The integration of diverse data sources and advanced feature selection methods led to the development of robust models. This study also emphasizes the significance of conducting multi-centric research and following the F.A.I.R. data principles to address challenges related to data heterogeneity. Future endeavors will focus on further refining these models, exploring harmonization techniques and addressing any potential privacy concerns in federated learning applications.

## Data Availability

Three public datasets –**HN1, HEAD-NECK and OPC**-were obtained from The Cancer Imaging Archive (TCIA) [35]. The **RADIOMICS-HN1** dataset [32] includes clinical data, volumetric CT, and PET scans from 137 patients with laryngeal carcinoma and oropharyngeal cancer (OPC), treated either with radiotherapy alone or with cisplatin or cetuximab. The **OPC-Radiomics** dataset [34] contains clinical data and CT scans of 606 OPC patients treated with either radiotherapy or chemo-radiotherapy. The **HEAD-NECK-PET-CT** dataset [33] consists of 298 patients with various HNC sub-sites, each with clinical descriptors, PET, and planning CT scans. The **RADIOMICS-HN3** dataset includes 165 patients with similar data characteristics to HN1. The **HCG** dataset comprises 370 patients treated with definitive chemo-radiation or external beam radiation using IGRT or IMRT alongside concurrent chemo-radiation. The **TMH** dataset includes 238 patients treated with SIB IM-IGRT and definitive radiotherapy or chemo-radiotherapy. At the time of this study, the **HN3, HCG,** and **TMH** datasets were not publicly available due to potentially identifiable patient information.

## Author contributions

VG was responsible for data processing, python coding, statistical experiments, and setting up the Vantage6 infrastructure. VG also prepared most of the manuscript text under the supervision of JvS and LW. BG adapted the feature selection and Cox models to the infrastructure and assisted VG with statistical experiments. JH helped VG with the dashboard development. GK and RS contributed to the infrastructure setup at CDAC, and UBS, AKJ, and SM helped with the infrastructure setup at TMH. KS and GL are the clinicians at HCG, SGL and SS are the clinicians at TMH and FJH is the senior clinician at Maastro. The clinicians helped the analysis by providing the data dictionary, and checking for clinical relevance of the models. GS, VR, AD, JvS, and LW jointly oversaw the overall supervision of the project. All authors reviewed the manuscript prior to submission and consented to its publication.

## Additional Information

### Ethics approval and consent to participate

Approval for re-use of standard-of-care (non-investigational) observational patient data in each partner institution (MAASTRO Clinic, Healthcare Global (HCG), and Tata Memorial Hospital (TMH)) was granted by their respective ethics review boards - waiving patient informed consent for the TRAIN retrospective observational study protocol (ClinicalTrials.gov Identifier NCT04655469).

All methods were performed in accordance with the relevant guidelines and regulations.

### Competing interests

The authors declare that they have no competing interests.

### Funding

VG, AD and LW acknowledge financial support from the Dutch Research Council (NWO) (TRAIN project, dossier 629.002.212) and the Hanarth Foundation.

## Acknowledgements

None.

## Notes

### Competing Interest Statement

The authors have declared no competing interest.

### Author Declarations

Approval for re-use of standard-of-care non-investigational observational patient data in each partner institution, MAASTRO Clinic, Healthcare Global, and Tata Memorial Hospital was granted by their respective ethics review boards, waiving patient informed consent for the TRAIN retrospective observational study protocol, ClinicalTrials.gov Identifier NCT04655469.

