## Supplementary Materials for "Federated Prediction Models and External Validation for Radiotherapy Outcomes in Oropharyngeal Cancer using F.A.I.R. Clinical and Radiomics Data"

*Table S1. Clinical case-mix summary statistics from each of the six data nodes, only for Oropharyngeal cases (NA – Not Available).*

|  | **HN1**  **(Maastro, NL)** | **OPC**  **(PMC, CA)** | **HEAD-NECK**  **(Quebec, CA)** | **HN3**  **(Maastro, NL)** | **HCG**  **(Bengaluru, IN)** | **TMH**  **(Mumbai, IN)** |
| --- | --- | --- | --- | --- | --- | --- |
| **Sample size**  **(only oropharyngeal)** | 88 | 606 | 203 | 63 | 108 | 63 |
| **Age in years**  Mean  Range | 59.8  44-80 | 60.5  33-89 | 62.9  34-90 | 61  29-78 | 59.8  32-83 | 58  29-87 |
| **Sex**  Female  Male | 21  67 | 125  481 | 52  151 | 16  47 | 17  91 | 9  54 |
| **Tumor stage**  T0  T1  T2  T3  T4  Tx | NA  14  28  13  33  NA | NA  103  198  183  122  NA | NA  26  91  51  34  1 | NA  9  18  12  24  NA | NA  5  25  47  31  NA | NA  17  20  16  10  NA |
| **Nodal stage**  N0  N1  N2  N3  Nx | 23  14  49  2  NA | 101  61  397  47  NA | 32  20  138  13  NA | 11  12  35  5  NA | 28  14  59  7  NA | 23  10  17  13  NA |
| **Metastasis stage**  M0  M1  Mx | 88  NA  NA | 606  NA  NA | 202  0  1 | 63  0  NA | 108  NA  NA | 63  NA  NA |
| **Overall stage (7^th^ ed.)**  I  II  III  IV | 6  8  14  60 | 11  38  85  472 | 2  15  27  159 | NA  NA  NA  NA | NA  NA  NA  NA | NA  NA  NA  NA |
| **HPV status**  Positive  Negative  Unknown | 23  57  8 | 356  143  107 | 76  19  108 | 34  29  NA | 13  45  50 | 10  28  25 |
| **Treatment**  Radiotherapy  Chemoradiotherapy | 51  37 | 309  297 | 48  175 | 29  34 | 7  101 | 19  44 |
| **Survival status**  Censored  Deceased | 38  50 | 347  259 | 179  24 | 36  27 | 87  21 | 44  19 |
| **Outcomes (in days)**  Mean survival time  Death events  Mean DM time  DM events  Mean LRR time  LRR events | 2251  50  1436  7  1369  18 | 2062  259  1995  83  1999  83 | 1364  24  1328  19  1315  20 | 1320  27  1288  8  1288  8 | 874  21  774  21  771  17 | 743  19  692  12  647  20 |
| **Radiomics extracted**  **(Sample size)** | 88 | 597 | 202 | 61 | 108 | 58 |

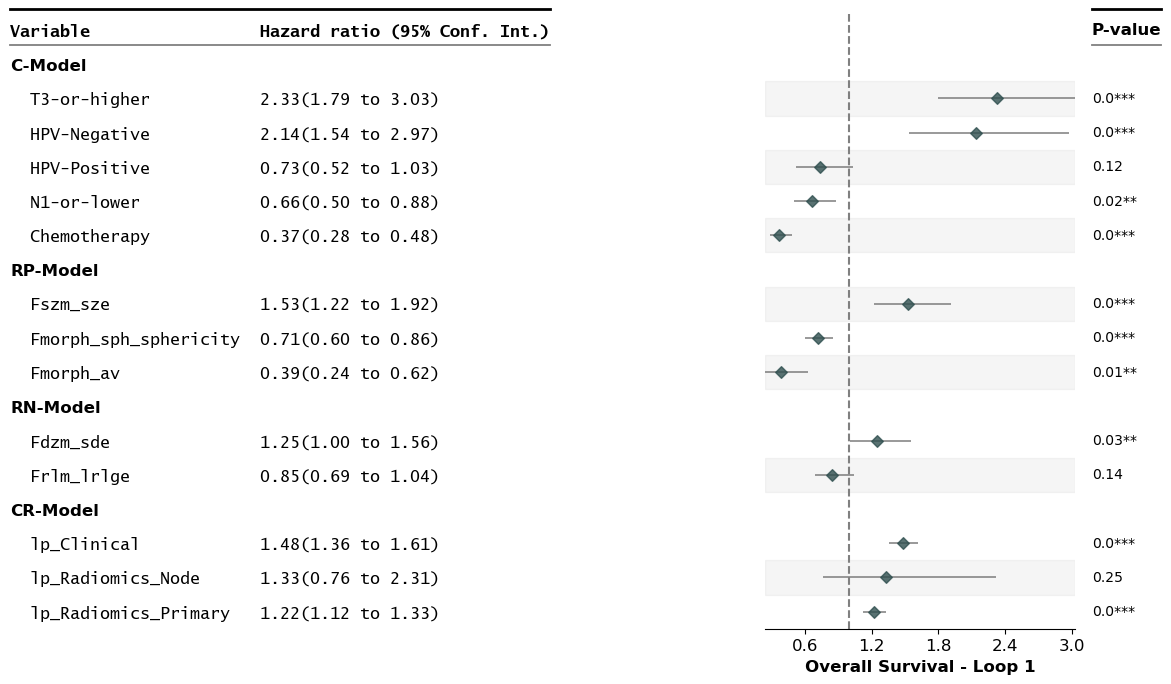

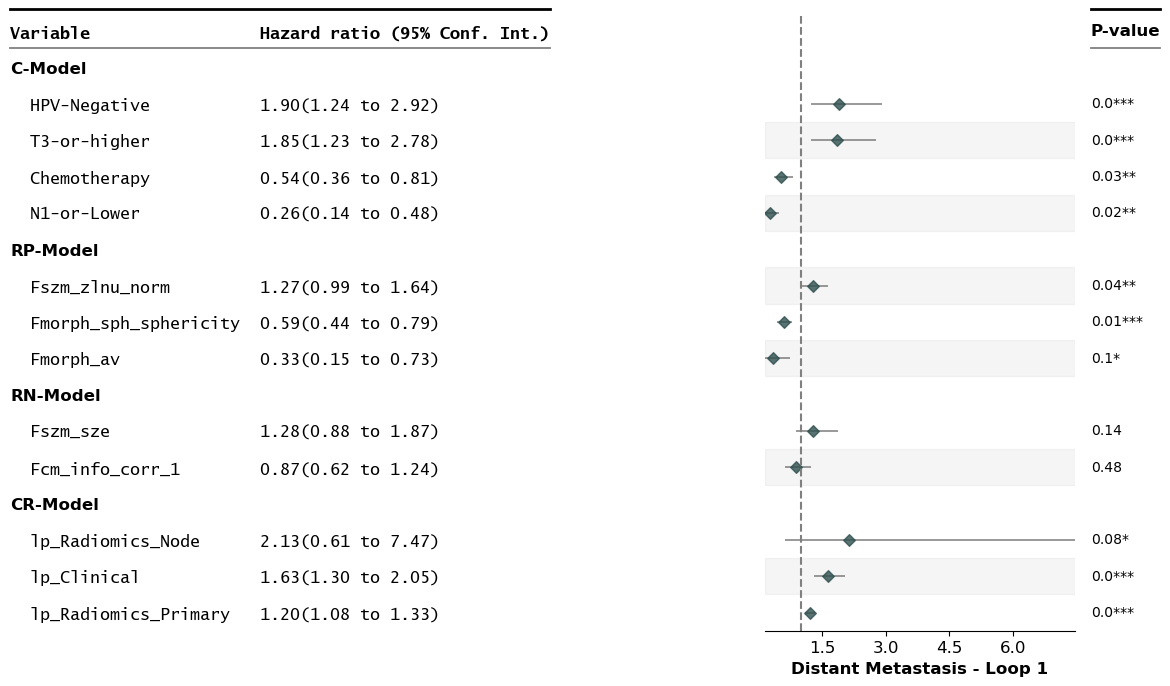

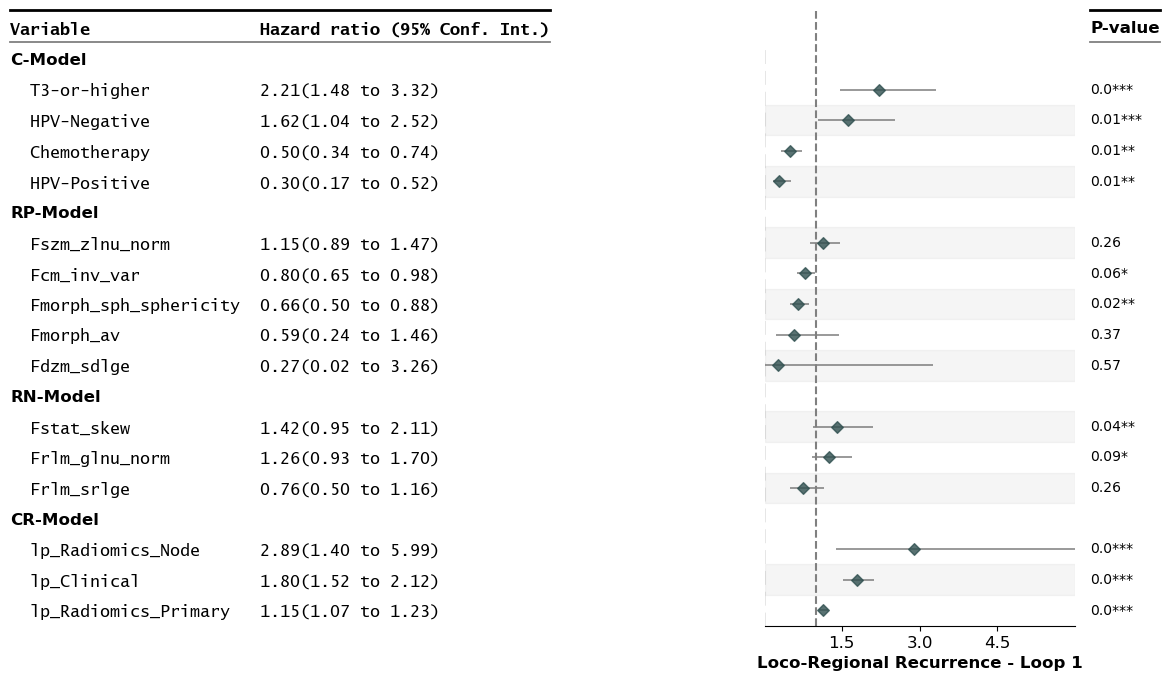

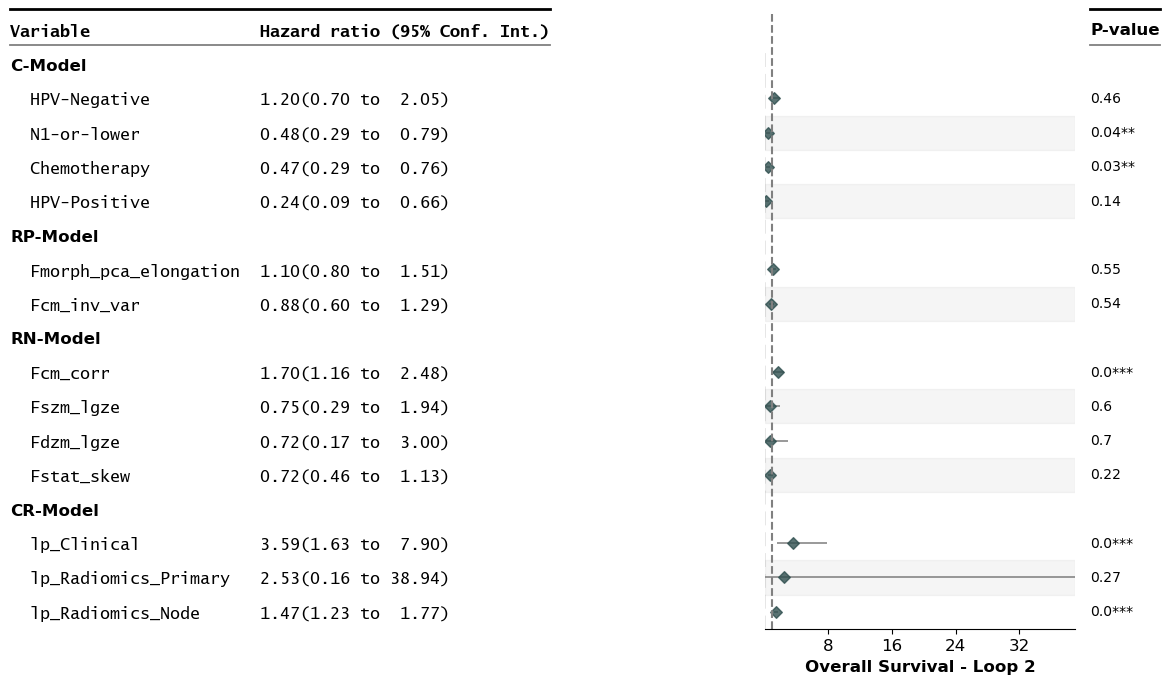

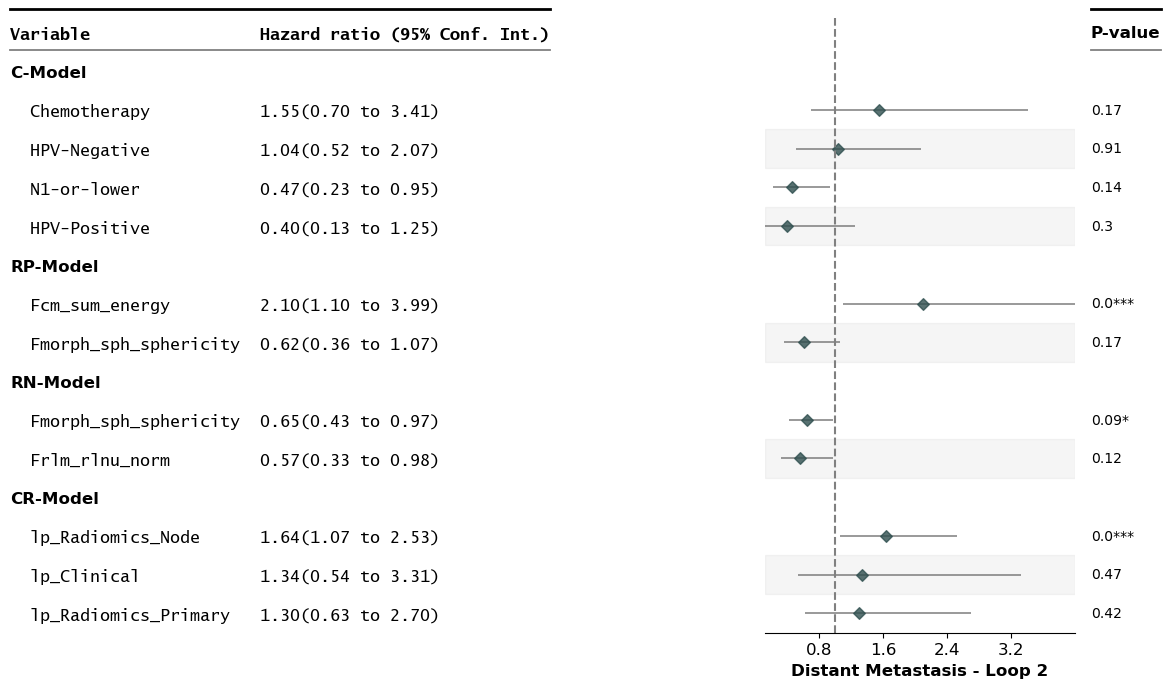

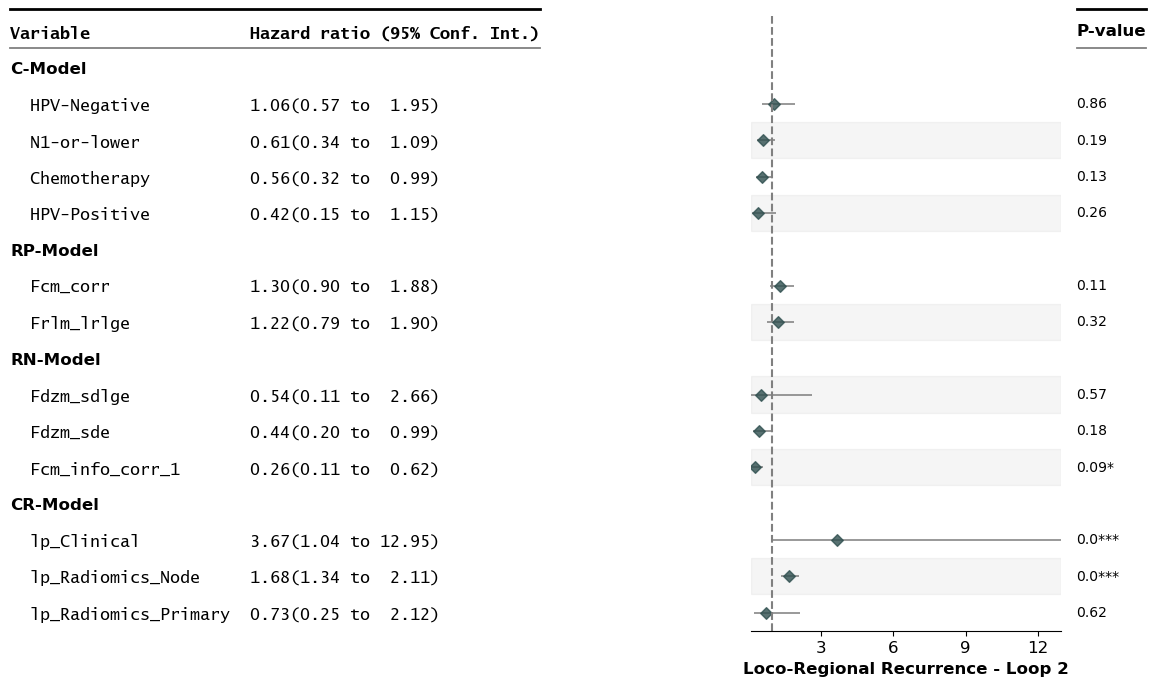

*Fig S1. Multi-variate federated CoxPH for OS, DM and LRR in two separate loops. Results shown with hazard ratios (HR) and 95% confidence intervals of the HR for the selected predictors. Models include C-Model (Clinical model), RP-Model (Radiomics Primary GTV Model), RN-Model (Radiomics Nodal GTV Model) and CR-Model (Combined model).*

*Table S2: C-index values for each validation cohort in loop 1, categorized by different outcomes.*

| **Training loop 1**  **(External validation results - C Index)** | **OS** | **DM** | **LRR** |
| --- | --- | --- | --- |
| **C-Model** |  |  |  |
| HN3 | 0.62 | 0.71 | 0.45 |
| HCG | 0.58 | 0.57 | 0.49 |
| TMH | 0.64 | 0.65 | 0.47 |
| **Global average** | **0.62** | **0.64** | **0.47** |
| **RP-Model** |  |  |  |
| HN3 | 0.56 | 0.63 | 0.61 |
| HCG | 0.47 | 0.50 | 0.55 |
| TMH | 0.55 | 0.54 | 0.57 |
| **Global average** | **0.53** | **0.55** | **0.57** |
| **RN-Model** |  |  |  |
| HN3 | 0.48 | 0.57 | 0.61 |
| HCG | 0.70 | 0.34 | 0.65 |
| TMH | 0.62 | 0.64 | 0.64 |
| **Global average** | **0.60** | **0.52** | **0.64** |
| **CR-Model** |  |  |  |
| HN3 | 0.63 | 0.69 | 0.53 |
| HCG | 0.43 | 0.47 | 0.60 |
| TMH | 0.58 | 0.54 | 0.59 |
| **Global average** | **0.55** | **0.56** | **0.57** |

*Table S3: C-index values for each validation cohort in loop 2, categorized by different outcomes.*

| **Training loop 2**  **(External validation results – C Index)** | **OS** | **DM** | **LRR** |
| --- | --- | --- | --- |
| **C-Model** |  |  |  |
| Mon | 0.59 | 0.58 | 0.63 |
| OPC | 0.68 | 0.59 | 0.74 |
| HN3 | 0.61 | 0.73 | 0.48 |
| **Global average** | **0.62** | **0.63** | **0.62** |
| **RP-Model** |  |  |  |
| Mon | 0.57 | 0.53 | 0.45 |
| OPC | 0.54 | 0.52 | 0.56 |
| HN3 | 0.46 | 0.61 | 0.60 |
| **Global average** | **0.52** | **0.55** | **0.54** |
| **RN-Model** |  |  |  |
| Mon | 0.52 | 0.58 | 0.71 |
| OPC | 0.48 | 0.51 | 0.55 |
| HN3 | 0.51 | 0.77 | 0.55 |
| **Global average** | **0.50** | **0.62** | **0.60** |
| **CR-Model** |  |  |  |
| Mon | 0.48 | 0.40 | 0.70 |
| OPC | 0.66 | 0.55 | 0.72 |
| HN3 | 0.56 | 0.81 | 0.51 |
| **Global average** | **0.57** | **0.59** | **0.64** |

*OS - Overall Survival, LRR – Loco Regional Recurrence, DM - Distant Metastasis, C-Model - Clinical Model, RP-Model - Radiomics Primary GTV Model, RN-Model - Radiomics Nodal GTV Model, CR-Model - Combined Model*

**PyRadiomics (v2.1.0) parameters:**

imageType:

Original:

binWidth: 25

featureClass:

shape: # Remove VoxelVolume, correlated to Volume

- Elongation

- Flatness

- LeastAxisLength

- MajorAxisLength

- Maximum2DDiameterColumn

- Maximum2DDiameterRow

- Maximum2DDiameterSlice

- Maximum3DDiameter

- MeshVolume

- MinorAxisLength

- Sphericity

- SurfaceArea

- SurfaceVolumeRatio

firstorder: # Remove Total Energy, correlated to Energy (due to resampling enabled)

- 10Percentile

- 90Percentile

- Energy

- Entropy

- InterquartileRange

- Kurtosis

- Maximum

- Mean

- MeanAbsoluteDeviation

- Median

- Minimum

- Range

- RobustMeanAbsoluteDeviation

- RootMeanSquared

- Skewness

- Uniformity

- Variance

glcm: # Disable SumAverage by specifying all other GLCM features available

- 'Autocorrelation'

- 'JointAverage'

- 'ClusterProminence'

- 'ClusterShade'

- 'ClusterTendency'

- 'Contrast'

- 'Correlation'

- 'DifferenceAverage'

- 'DifferenceEntropy'

- 'DifferenceVariance'

- 'JointEnergy'

- 'JointEntropy'

- 'Imc1'

- 'Imc2'

- 'Idm'

- 'Idmn'

- 'Id'

- 'Idn'

- 'InverseVariance'

- 'MaximumProbability'

- 'SumEntropy'

- 'SumSquares'

glrlm:

glszm:

gldm:

ngtdm:

setting:

interpolator: 'sitkBSpline'

resampledPixelSpacing: [2, 2, 2]

padDistance: 10 #Extra padding for large sigma valued LoG filtered images

resegmentRange: [-3, 3]

resegmentMode: sigma

voxelArrayShift: 1000

label: 1

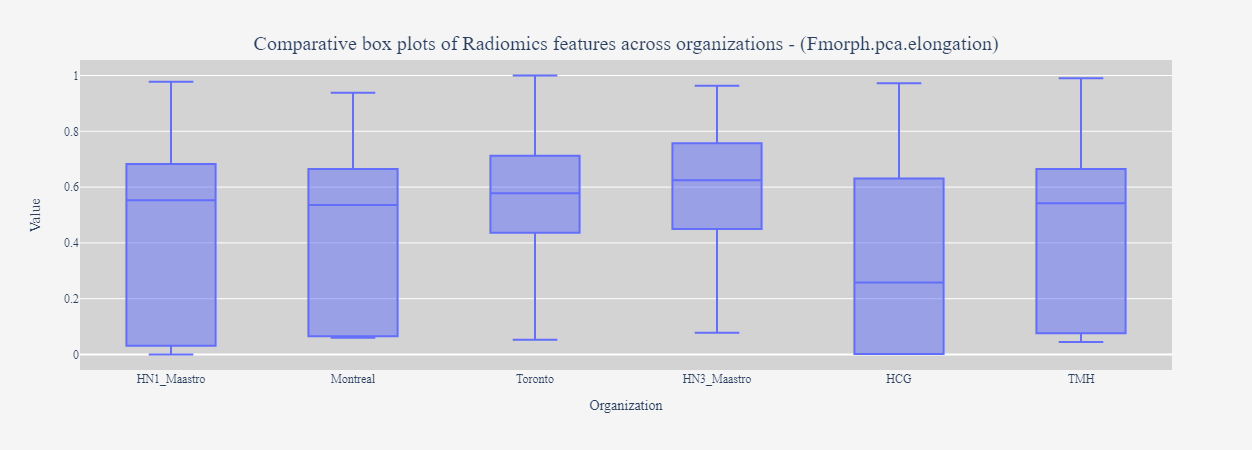

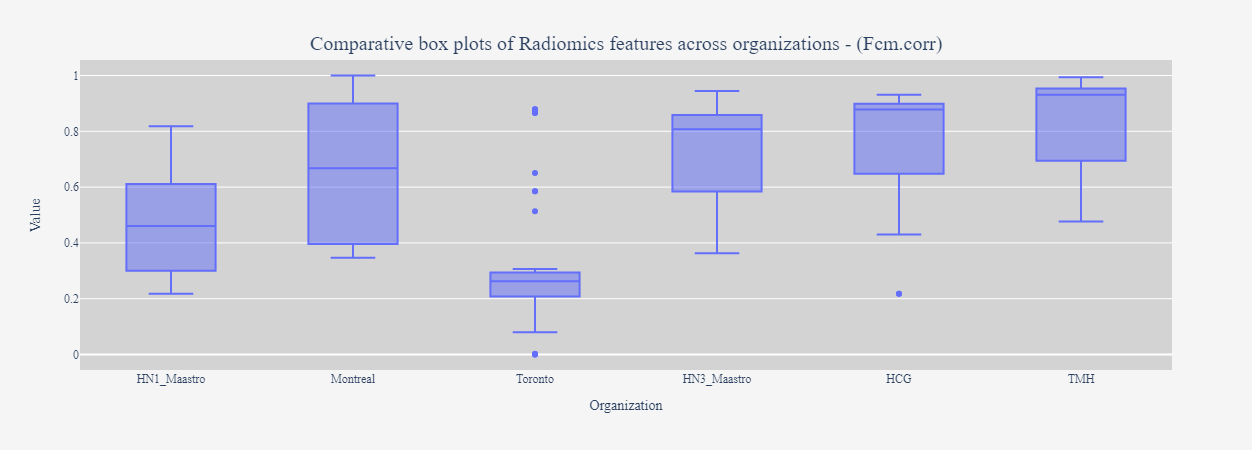

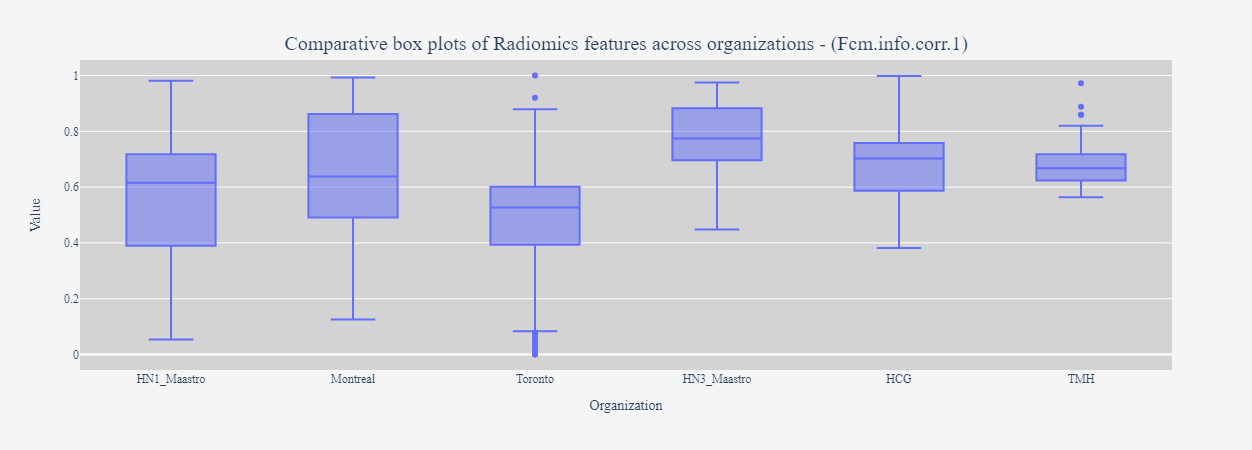

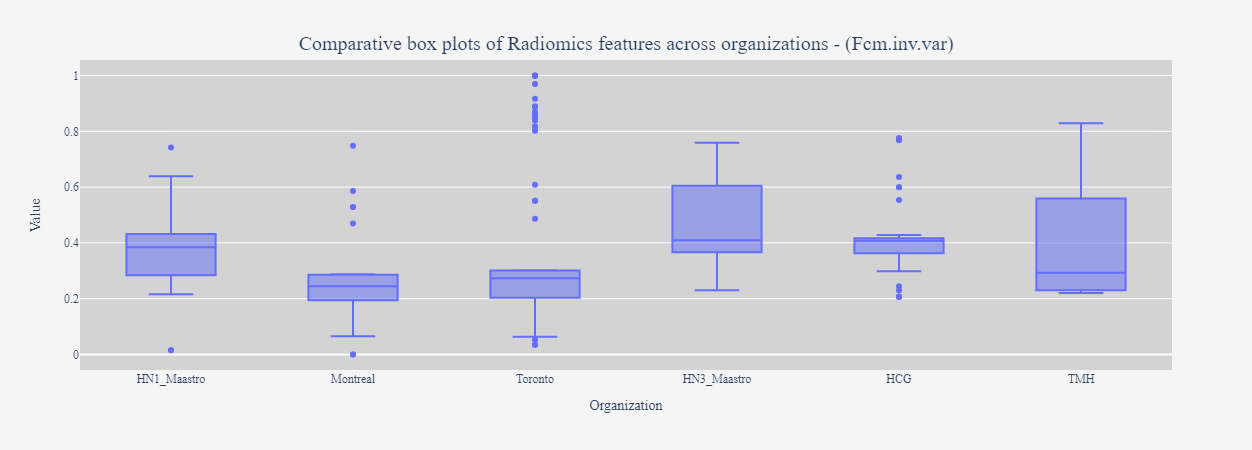

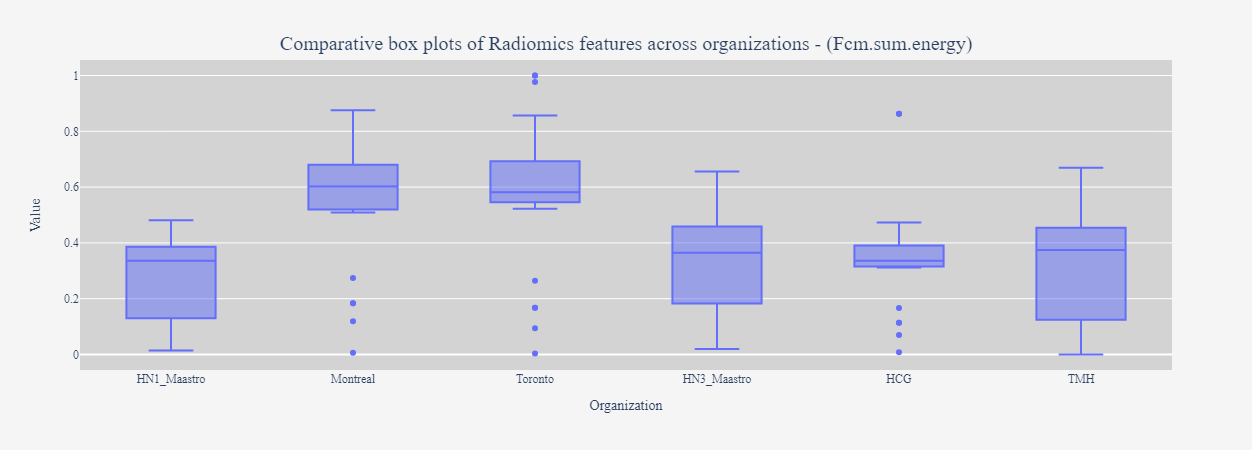

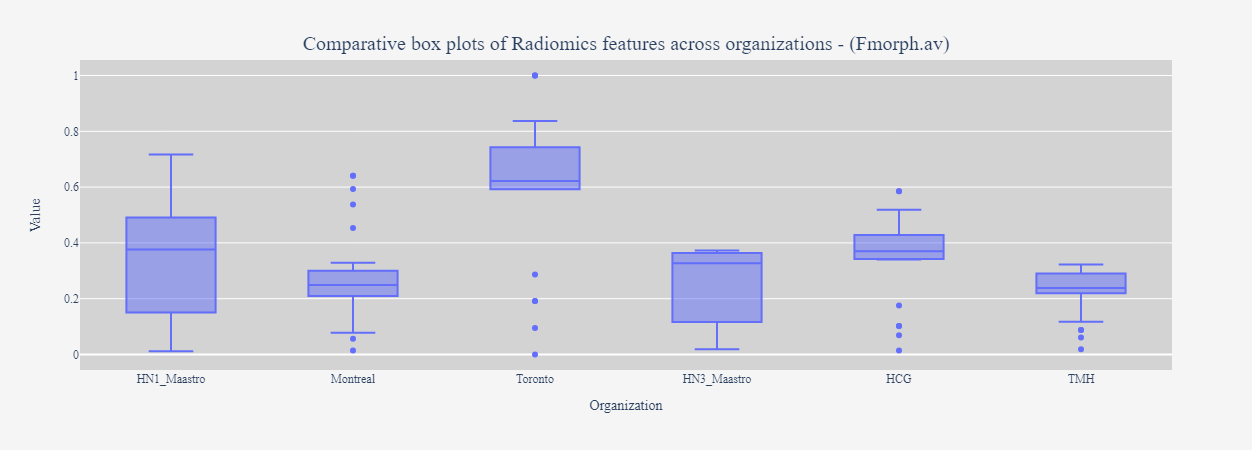

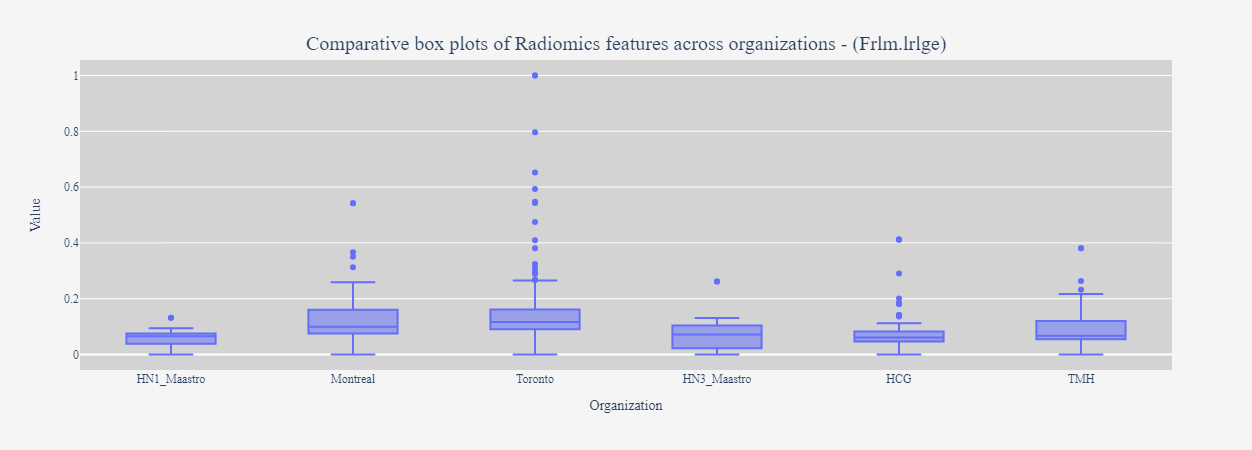

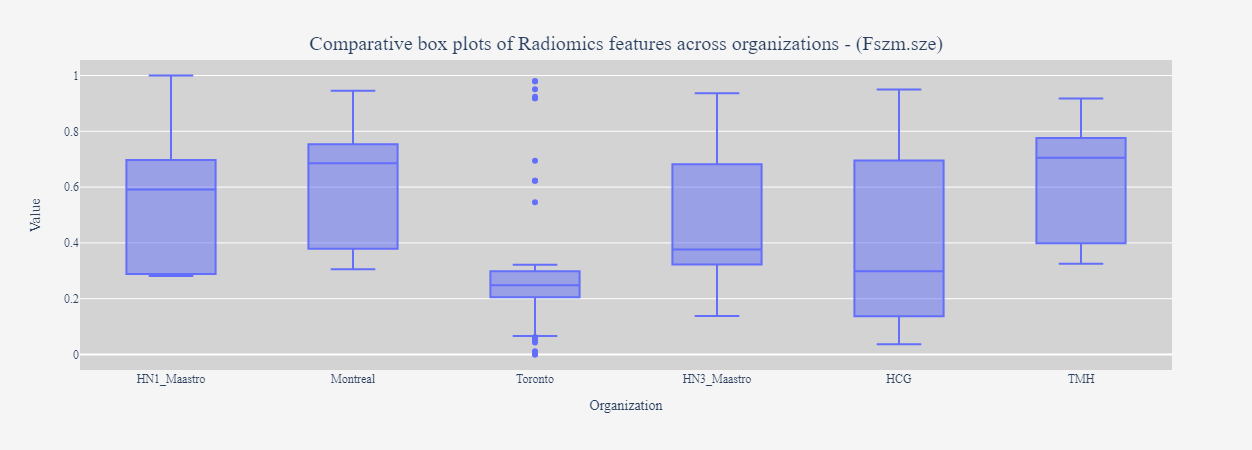

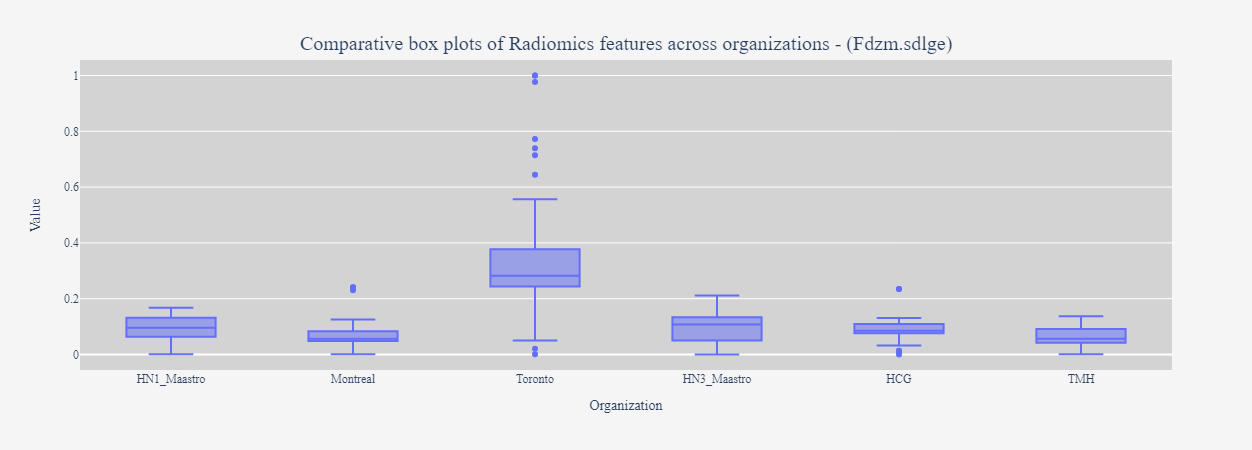

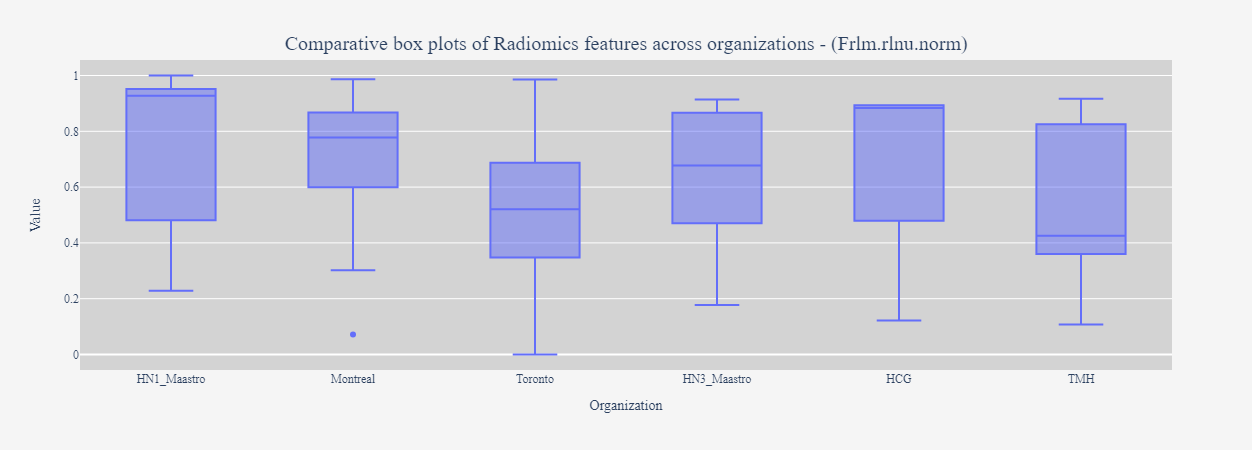

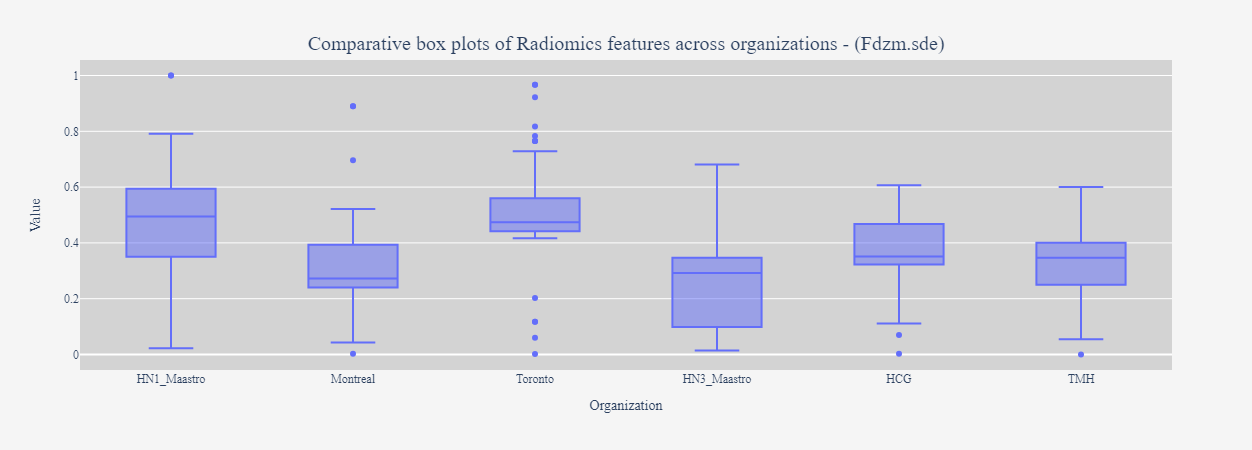

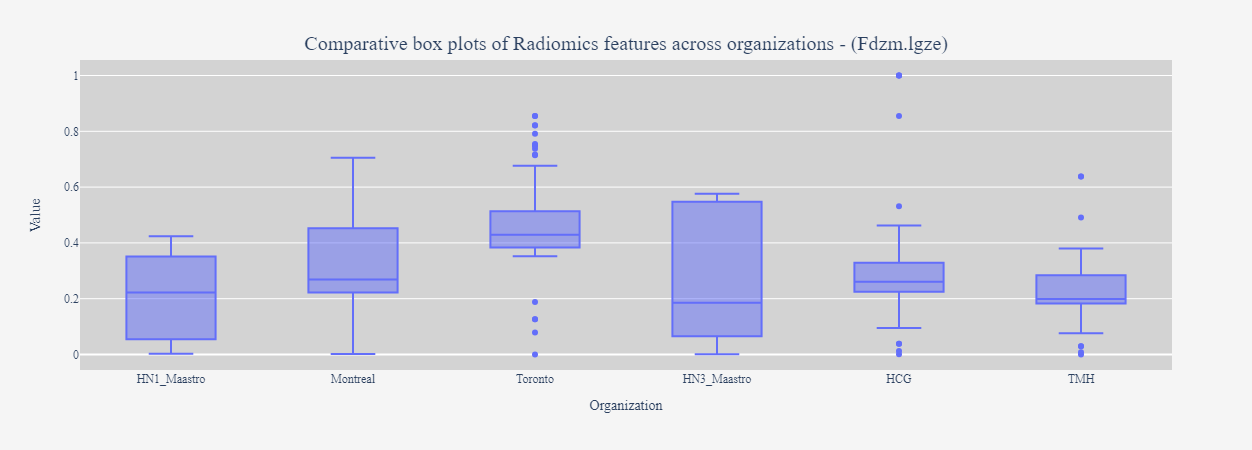

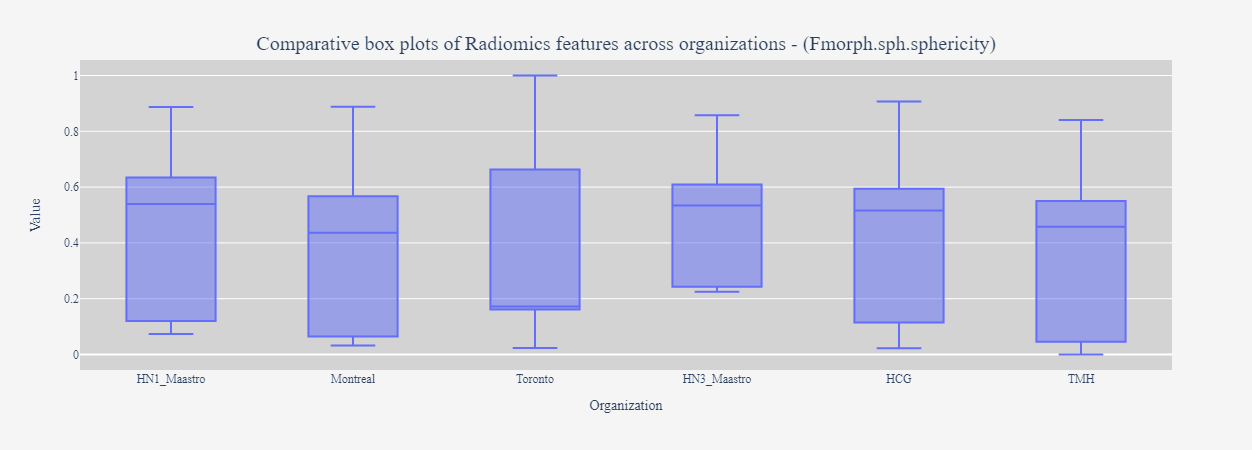

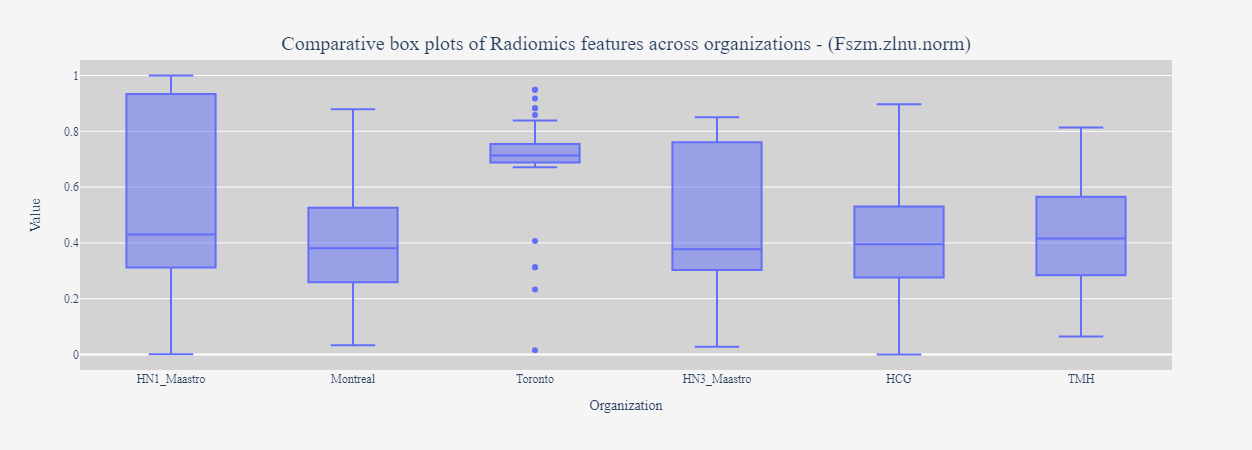

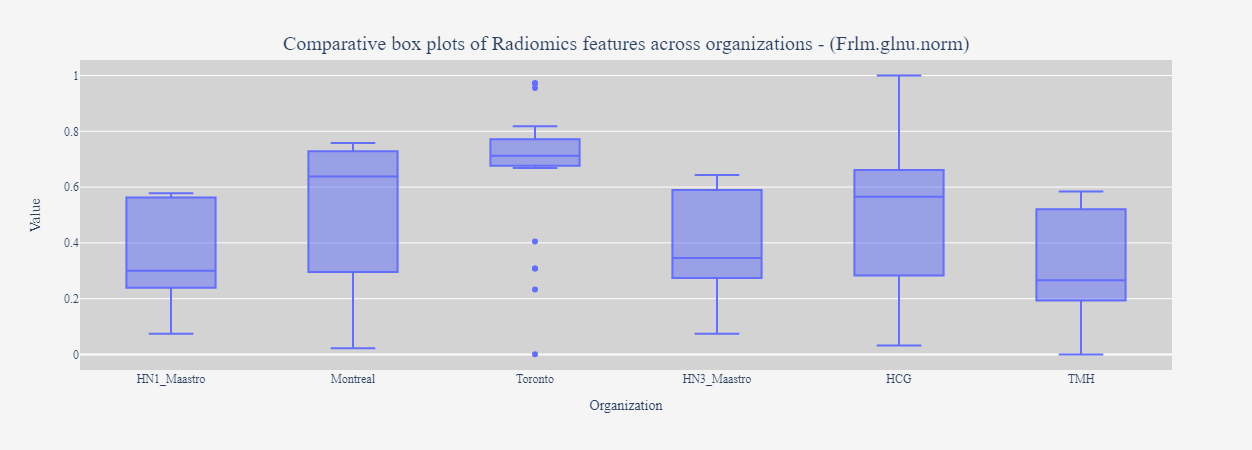

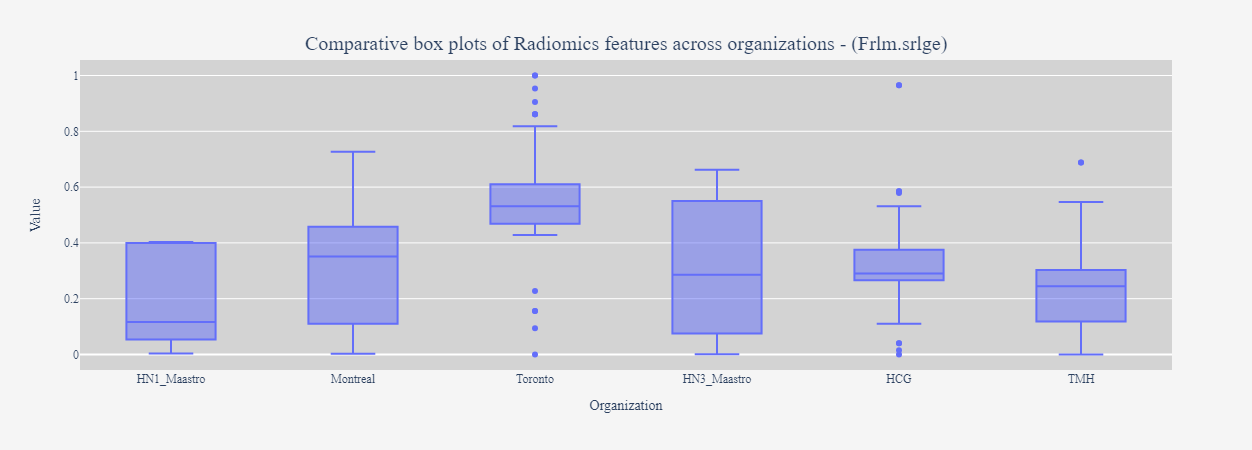

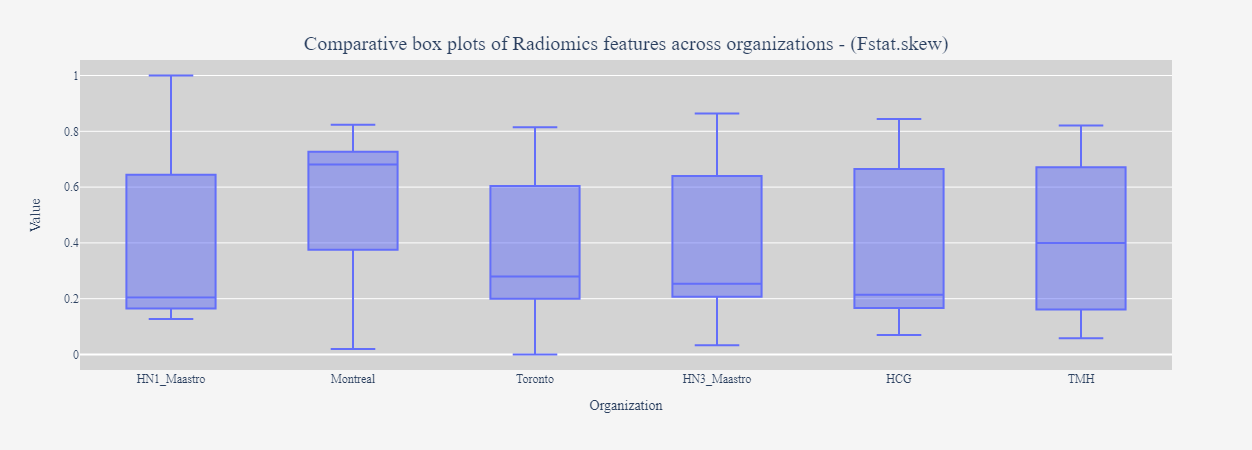

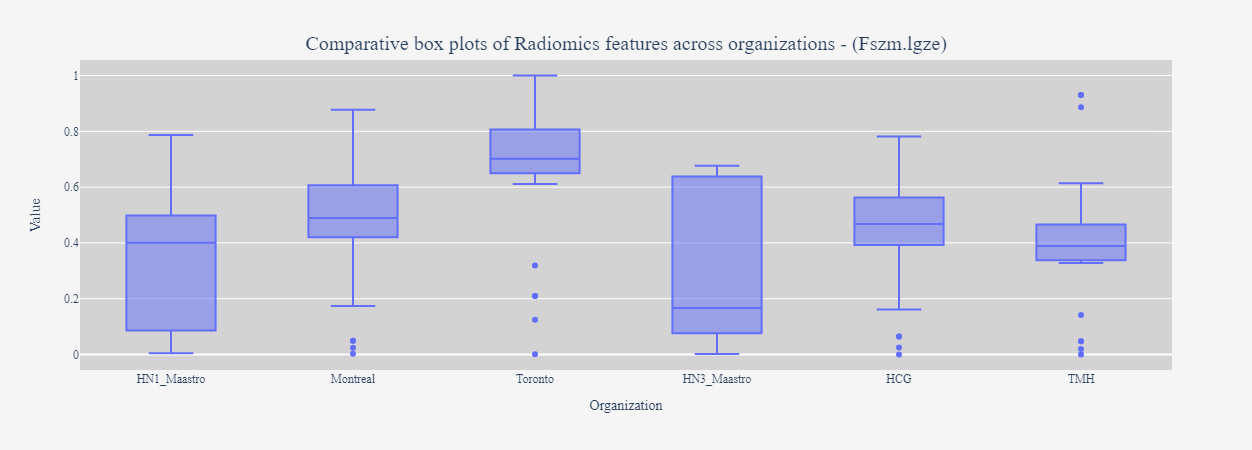

*Fig S2: Comparative box blots for the selected radiomics features across six datasets*

**Project reproducibility requirements**

Workflow package for RDF conversion: Flyover (project branch: flyover-DCR)

Workflow home page: <https://github.com/MaastrichtU-CDS/Flyover>

Modeling and dashboard scripts: <https://github.com/MaastrichtU-CDS/TRAIN-DCR-tools>

Operating system: Platform-independent

Recommended processor, memory, and storage: Intel i5 – 8^th^ gen equivalent or higher, 8GB RAM, at least 16GB storage.

Programming language: Python and Java

Other requirements: Either Docker Desktop (Windows/MacOS) or Docker Engine (Linux), Docker Compose 3.7.

License: Docker Desktop/Engine is licensed as part of either a free or paid Docker subscription (<https://www.docker.com/pricing/faq/>). Jupyter/base-notebook image is distributed under a Modified BSD License. GraphDB Free Edition is distributed as freeware.

Restrictions on use by non-academics: Creative Commons By Attribution Non Commercial only version 4.0 International (CC BY-NC 4.0).

**List of abbreviations**

1. C-index: Concordance Index

2. CFS: Correlation-based Feature Selection

3. CoxPH: Cox Proportional Hazards

4. CT: Computed (x-ray) tomography

5. DICOM: Digital Imaging and Communications in Medicine

6. DM: Distant Metastasis

7. F.A.I.R: Findable Accessible Interoperable Reusable

8. GTV: Gross Tumor Volume

9. H&N: Head and Neck

10. HR: Hazard Ratio

11. IBSI: The Image Biomarker Standardization

12. LASSO: Least Absolute Shrinkage and Selection Operator

13. LOOCV: Leave-One-Out Cross-Validation

14. LRR: Loco-regional Recurrence

15. OMOP-CDM: Observational Medical Outcomes Partnership - Common Data Model

16. OS: Overall Survival

17. OWL: Web Ontology Language

18. PHT: Personal Health Train

19. RDF: Resource Description Framework

20. RDFS: Resource Description Framework Schema

21. RTSTRUCT: Radiotherapy Structure Sets

22. SOP: Service Object Pair

23. SPARQL: SPARQL Protocol and RDF Query Language

24. TCIA: The Cancer Imaging Archive

25. UID: Unique Identifiers
